# Digital breast tomosynthesis(DBT) vs 2D mammography and impact of combined use: A meta-analysis

**DOI:** 10.1101/2023.12.07.23299674

**Authors:** Dev Andharia, Hetvi Shah, Anushka Dipakkumar Prajapati, Aayushi Darshan Bhansali, Abhijay B. Shah, Dev Desai

## Abstract

**Background:** Having breast cancer and receiving treatment for it is viewed as a very traumatic experience for women. From the recent advances in breast cancer diagnosis protocol; The triple assessment for a lump in the breast is the best practice and its application towards early disease detection is crucial. For which, 2D mammography is used. The limitation of this technique is a potential tissue overlap in dense breasts. Digital breast tomosynthesis (DBT) is a new imaging technology that can address the limitations of 2D mammography. This study aims to highlight the use of DBT.

**Methods:** The study population included all women aged from 18 to 80. For this analysis, the subdivision of the population is not created. A total of 28 RCTs with a total of 1,735,126 patients were selected for the DBT+FFDM vs FFDM study and 5 RCTs with a total of 554,419 patients were selected for the DBT vs FFDM study.

**Results:** For the DBT+FFDM vs FFDM; The overall diagnostic odds ratio was 1.28 with a 95%CI [1.22-1.33], the Decrease in RR is 2.43% and the increase in CDR is 1.583. for DBT vs FFDM; the overall diagnostic odds ratio was 1.38 with a 95%CI [1.28-1.50], a decrease in RR is 1.70%, and an increase in CDR is 1.4.

**Conclusion:** DBT+FFDM and DBT do outweigh FFDM alone.

## Introduction

Breast cancer is the most common cancer observed in females overall. Having breast cancer and receiving treatment for it is viewed as a very traumatic experience for women because this reflects on their self-image, sexual relationship, and self-confidence and this may lead to other psychological problems such as anxiety, denial, depression, anger, etc.[1] This is why early breast cancer detection with minimum recalls is useful to avoid fears and anxiety toward the disease.[2]

From the recent advances in breast cancer diagnosis protocol; The triple assessment for a lump in the breast is the best practice and its application towards early disease detection is crucial.[3] For screening of breast cancer, there are two main resources available; Mammography and a second clinical breast examination.[4]

For mammography, there are few options available, Conventional two-dimensional (2D) mammography is widely acknowledged as the most effective method for detecting breast cancer. A meta-analysis of 11 randomized trials concluded that mammography screening can lead to a reduction of 20% mortality in breast cancer.[5]

The limitation of this technique is a potential tissue overlap in dense breasts. This can consequently obscure the area of interest in the image, which can lead to false-negative and false-positive readings, following which recall of the patient for further imaging and biopsy will be needed. This explains 15-30% of undetected cancers by standard screening protocol. This can in turn be responsible for anxiety and non-attendance for the next routine breast screening tests.[6]

Digital breast tomosynthesis (DBT) is a new imaging technology that can address the limitation caused by overlapping structures in 2D mammography. This technique provides a dual benefit for screening purposes. First, DBT increases the cancer detection rate mostly by highlighting architectural distortions [AD] and allowing a better assessment of the shape and margins of the mass. Second, it helps reduce the recall rate. However, DBT is not included in the majority of cancer screening programs worldwide.[5], [7], [8]

The use of DBT with digital or synthetic mammography for breast cancer screenings increases the rates of overall and invasive breast cancer detection. however, no evidence shows that DBT, compared with digital or synthetic mammography may decrease recall rates, with high or moderate quality.[9]

This study aims to combine different results and create the most accurate meta-analysis to date to experience clinical routine use of tomosynthesis, with an evaluation of the added value of this technique by comparing the diagnostic accuracy with the increase in cancer detection rate [CDR] and decreases in recall rate [RR] of Digital breast tomosynthesis vs 2D mammography and DBT combined with 2D mammography vs the accuracy of 2D mammography alone.

From here onwards, Digital breast tomosynthesis [DBT] means 3D mammography and conventional mammography means 2D mammography; Full-Field Digital Mammography[FFDM]; Digital mammography[DM] or standard 2D mammography. These words are used interchangeably.

**Image 1:**
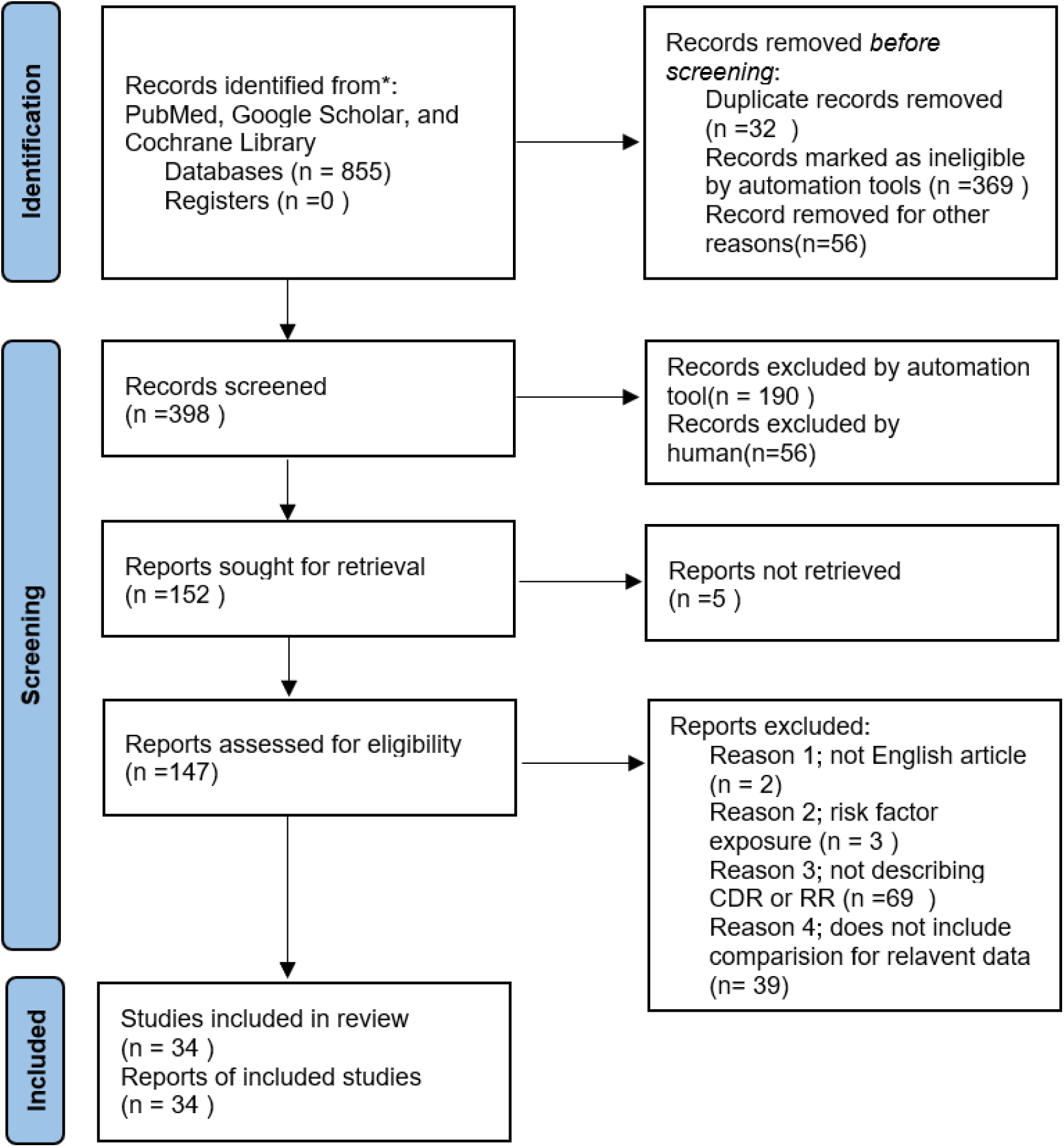
PRISMA Flow chart.

## METHODOLOGY

### DATA COLLECTION

For the data collection, Researchers reviewed all relevant literature, a search was done using PubMed, Google Scholar, and Cochrane Library databases.

The used medical subject headings (MeSH) and keywords were the following: ‘3D vs 2D mammography’, ‘DBT+FFDM vs FFDM’, ‘DBT vs FFDM’, ‘Digital Breast Tomosynthesis-meta analysis’, ‘DBT vs DM’, ‘DBT vs 2d mammography’, and ‘3d and 2d mammography’.

For further data collection from all the articles, a review of all the references and meta-analyses available, titles and abstracts were read methodically, and papers with detailed information about the study group and control group were selected.

### INCLUSION AND EXCLUSION CRITERIA

In this paper, the data is compiled for the studies that provided Recall rates and cancer detection rates in the women who were not exposed to the risk factors.

The main purpose of this study is to compare and provide the decrease in Recall rate and increase in cancer detection rate overall for the implementation of the DBT+FFDM method in routine screening protocol, The study population includes all women aged from 18 to up to 80. The subdivision of the population was not created as it is beyond the scope of this study. The following papers were excluded; Carbonaro et al 2016 [10], Michell et al 2012[11], and Chae et al 2016[12] as the study populations were exposed to risk factors thus hindering the result analysis.

Inclusion criteria were the following: 1) only articles in the English language were selected, 2) Articles providing DBT vs FFDM comparison. 3) Articles with DBT+FFDM vs FFDM comparison 4) articles providing either of the following cancer detection rate [CDR] and recall rate [RR].

Exclusion criteria were the following: 1) non-English language articles, 2) articles where the study group was exposed to the risk factors 3) articles where provisional diagnoses for the patient were given as architectural distortion. 4) articles not providing CDR or RR.

### DATA EXTRACTION

According to PRISMA, a total of 28 RCTs with a total of 1,735,126 patients were selected for the DBT+FFDM vs FFDM study and 5 RCTs with a total of 554,419 patients were selected for the DBT vs FFDM study. Sumkin et al 2015 were only included in the RR table. Every appearing paper was independently studied by three different reviewers. Each article was analyzed for the number of patients, age, procedure modality, risk factors, BIRADs scoring of patients, recall rates, and cancer detection rate. Further discussion and consultation with the other authors and third-party reviewers were used to resolve conflicts. In the majority of the papers, the recall rate (RR) was calculated as the number of positive examinations (BI-RADS 0, 3, and 4) over the total number of screening examinations interpreted while Some papers considered only BI-RADS cat 0 for the calculation of RR. And some also include Cat 5. The modified Jadad score was employed to establish the quality of each paper for participation.

### ASSESSMENT OF STUDY QUALITY

Three writers independently assessed the caliber of each included study. This test consists of 10 questions, each with a score between 0 and 2, with 20 being the maximum possible overall score. Two authors rated each article independently. For deciding the bias risk for RCTs, the Cochrane tool was applied. There were no assumptions made for any lacking or unclear material. There was no funding involved in collecting and reviewing the data analysis.

### STATISTICAL ANALYSIS

For the determination of the odds ratio and other analyses, The statistical software packages RevMan (Review Manager, version 5.3), SPSS (Statistical Package for the Social Sciences, version 20), and Excel were used. Fixed- and random-effects models were used to estimate diagnostic odds ratios (DOR) with 95 percent confidence intervals to examine critical clinical outcomes. The odds ratio of the individual study was plotted on the funnel plot and forest chart.

### BIAS STUDY

The risk of bias was evaluated by the QUADAS-2 analysis. which includes 4 Major parameters, as follows; 1. Patient selection, 2. Index test, 3. Reference standard, 4. The patient flow and Timing of the Index tests.

## RESULT

**Table 1:** Table of the description of papers.

| Author Name and Year | Location | Duration of the study | Population | Study design | Modalities | Increase in CDR |  | Decrease in Recall rate |  |
| --- | --- | --- | --- | --- | --- | --- | --- | --- | --- |
|  |  |  |  |  |  | DBT | DBT+DM | DBT | DBT+DM |

|  |  |  |  |  |  | v DM | v DM | v DM | v DM |
| --- | --- | --- | --- | --- | --- | --- | --- | --- | --- |
| Alsheik et al 2019[13] | US | Jun 2015-Sept 2017 | 99660 (DM)<br>95370 (DBT) | Prospective study | DBT vs DM | 1 |  | -2.15 |  |
| Aujero et al 2017[14] | US | Oct 2011-Jun 2016 | 32076 (DM)<br>30561 (DBT +DM) | Retrospective study | DBT+DM vs DM |  | 1.1 |  | -2.9 |
| Bahl et al 2018[15] | US | Jan 2009-Feb 2011 (DM)<br>Jan 2013-Feb 2015 (DBT) | 78 385 (DM)<br>76 896 (DBT +DM) | Retrospective study | DBT+DM vs DM |  | ResultA:<br>0<br>ResultB:<br>0 |  |  |
| Bernardi et al 2016[16] | Italy | May 2013-May 2015 | 9677 (DM)<br>9677 (DBT +DM) | Prospective study | DBT+DM vs DM |  | 2.2 |  | 0.55 |
| Chikarmane et al 2020[17] | US | Oct 2014-Sept 2016 (DM)<br>Feb 2017-Dec 2018 (DBT) | 5706 (DM)<br>4440 (DBT +DM) | Retrospective study | DBT+DM vs DM |  | 0.1 |  | -2.24 |
| Ciatto et al 2013[18] | Italy | Aug 2011-Jun 2012 | 7294 (DM)<br>7294 (DBT +DM) | Prospective comparison study | DBT+DM vs DM |  | 2.8 |  | -1 |
| Cohen et al 2018[19] | US | Feb 2011-Jun 2014 | 71656 (DM)<br>31414 (DBT +DM) | Retrospective review | DBT+DM vs DM |  | 0.8 |  | -1.8 |
| Conant et al 2016[20] | US | Jan 2011-Dec 2014 | 113061 (DM)<br>25268 (DBT +DM) | Retrospective analysis of prospective data | DBT+DM vs DM |  | 1.5 |  | -1.7 |
| Conant et al 2020[21] | US | Sept 2010-Sept 2016 | 10511 (DM)<br>56839 (DBT) | Retrospective study | DBT vs DM | 1 |  | -2.4 |  |
| Destounis et al 2014[22] | US | Jun 2011-Dec 2011 | 524 (DM)<br>524 (DBT +DM) | Retrospective review | DBT+DM vs DM |  | 1.9 |  | -7.25 |
| Durand et al 2015[23] | US | Aug 2011-Dec 2012 | 9364 (DM)<br>8591 (DBT +DM) | Retrospective review | DBT+DM vs DM |  | 0.2 |  | -4.5 |
| Friedewald et al 2014[24] | US | Mar 2010-Dec 2012 | 2,80,698 (DM)<br>1,72,727 (DBT +DM) | Retrospective multi-centre analysis | DBT+DM vs DM |  | 1.2 |  | -1.63 |
| Giess et al 2017[25] | US | Oct 2012-May 2015 | 14180 (DM)<br>9817 (DBT +DM) | Retrospective study | DBT +DM vs DM |  | 2 |  | 0.4 |
| Greenberg et al 2014[26] | US | Aug 2011-Nov 2012 | 38674 (DM)<br>20943 (DBT +DM) | Retrospective multisite study | DBT+DM vs DM |  | 1.4 |  | -2.6 |
| Haas et al 2013[27] | US | Oct 2011-Sept 2012 | 7058 (DM)<br>6100 (DBT +DM) | Retrospective study | DBT+DM vs DM |  | 0.5 |  | -3.6 |
| Heindel et al 2022[28] | Germany | July 2018- Dec 2020 | 49762 (DM)<br>49 715 (DBT +DM) | Prospective RCT | DBT+DM vs DM |  | 2.3 |  |  |
| Houssami et al 2014[29] | Italy | Aug 2011- Jun 2012 | 7292 (DM)<br>7292 (DBT +DM) | Prospective study | DBT+DM vs DM |  | ResultA: 2.7<br>ResultB: 2.8 |  | ResultA: -0.9<br>ResultB: -1 |
| Houssami et al 2017[30] | Italy | May 2013- May 2015 | 9677 (DM)<br>9677 (DBT +DM) | Secondary analysis<br>Prospective study | DBT+DM vs DM |  | 1.9 |  |  |
| Lang et al 2016[31] | Sweden | Jan 2010- Dec 2012 | 7500 (DM)<br>7500 (DBT +DM) | Prospective study | DBT +DM vs DM |  | 2.6 |  | 1.2 |
| Lourenco et al 2015[32] | US | Mar 2011- Feb 2012 (DM)<br>Mar 2012- Feb 2013 (DBT) | 12577 (DM)<br>12921 (DBT) | Retrospective study | DBT vs DM | 0.4 |  | -2.94 |  |
| McCarthy et al 2014[33] | US | Sept 2010- Aug 2011 (DM)<br>Oct 2011- Feb 2013 (DBT +DM) | 10728 (DM)<br>15571 (DBT +DM) | Retrospective – Prospective study | DBT +DM vs DM |  | 0.9 |  | -1.6 |
| McDonald et al 2015[34] | US | Sept 2010- Aug 2011 (DM)<br>Oct 2011- Feb 2013 (DBT +DM) | 10728 (DM)<br>15571 (DBT +DM) | Retrospective study | DBT +DM vs DM |  | ResultA: 1.7<br>ResultB: 0.8 |  | ResultA: -4.5<br>ResultB: -1.3 |
| McDonald et al 2016[35] | US | Sept 2010- Sept 2014 | 10728 (DM)<br>33740 (DBT +DM) | Retrospective study | DBT +DM vs DM |  | ResultA: 1.5<br>ResultB: 1.2<br>ResultC: 0.9 |  | ResultA: -1.6<br>ResultB: -1.4<br>ResultC: -1.2 |
| Pattacini et al 2018[36] | Italy | Mar 2015- Mar 2016 | 9783 (DM)<br>9777 (DBT +DM) | Prospective RCT | DBT+DM vs DM |  | 4.1 |  | 0 |
| Powell et al 2017[37] | US | Jun 2012- Aug 2014 | 10477 (DM)<br>2304 (DBT +DM) | Retrospective study | DBT +DM vs DM |  | 2.6 |  | -2.33 |
| Rose et al 2013[38] | US | May 2011- Jan 2012 | 13856 (DM)<br>9499 (DBT +DM) | Retrospective study | DBT+DM vs DM |  | 1.33 |  | -3.2 |
| Rose et al 2014[39] | US | May 2011- Jan 2012 | 10878 (DM)<br>10878 (DBT +DM) | Prospective interpretations of DBT compared with Retrospective readings of DM | DBT+DM vs DM |  | 1.9 |  | -2.8 |
| Roth et al 2014[40] | US | Sept 2010- | 10751 (DM) | Interval analysis | DBT+DM vs DM |  | 1 |  | -1.6 |
|  |  | Aug 2011 (DM)<br>Sept 2011-Aug 2012 (DBT+DM) | 11115 (DBT +DM) |  |  |  |  |  |  |
| Sharpe et al 2016[41] | US | Jan 2011-Mar 2014 | 80149 (DM)<br>5703 (DBT +DM) | Prospective study with Retrospective cohort | DBT +DM vs DM |  | 1.9 |  | -1.41 |
| Skaane et al 2013[42] | Norway | Nov 2010-Dec 2011 | 12621 (DM)<br>12621 (DBT +DM) | Prospective study | DBT+DM vs DM |  | 2.3 |  | -0.77 |
| Starikov et al 2016[43] | US | Jan 2013-Dec 2013 | 12157 (DM)<br>2070 (DBT +DM) | Retrospective cohort | DBT+DM vs DM |  | 2.1 |  | -7.3 |
| Sumkin et al 2015 [44] | US | May 2010-Sept 2014 | 1074 (DM)<br>1074 (DBT +DM) | Prospective single-site clinical study | DBT+ DM vs DM |  |  |  | -12.9 |
| Winter et al 2020[45] | US | Jan 2012-Dec 2014 (DM)<br>Jan 2016-Dec 2018 (DBT) | 117099 (DM)<br>119746 (DBT) | Retrospective study | DBT vs DM | 2.4 |  | -2.1 |  |
| Zackrisson et al 2018[46] | Sweden | Jan 2010-Feb 2015 | 14848 (DM)<br>14848 (DBT) | Prospective survey | DBT vs DM | 2.2 |  | 1.1 |  |

### DBT+FFDM vs FFDM

A total of 28 RCTs with 1,735,126 patients were selected for the study of DBT+FFDM vs FFDM arm. In Image 2, the forest chart and in Image 3, funnel plots are described. This image suggests as the sample size increases, the odds ratio increases, which can be seen in Cohen et al 2019, Conant et al 2016, Friedewald et al 2014, Greenberg et al 2014, and Heindel et al 2022. The overall diagnostic odds ratio was 1.28 with a 95%CI [1.22 to 1.33].

**Image 2:**
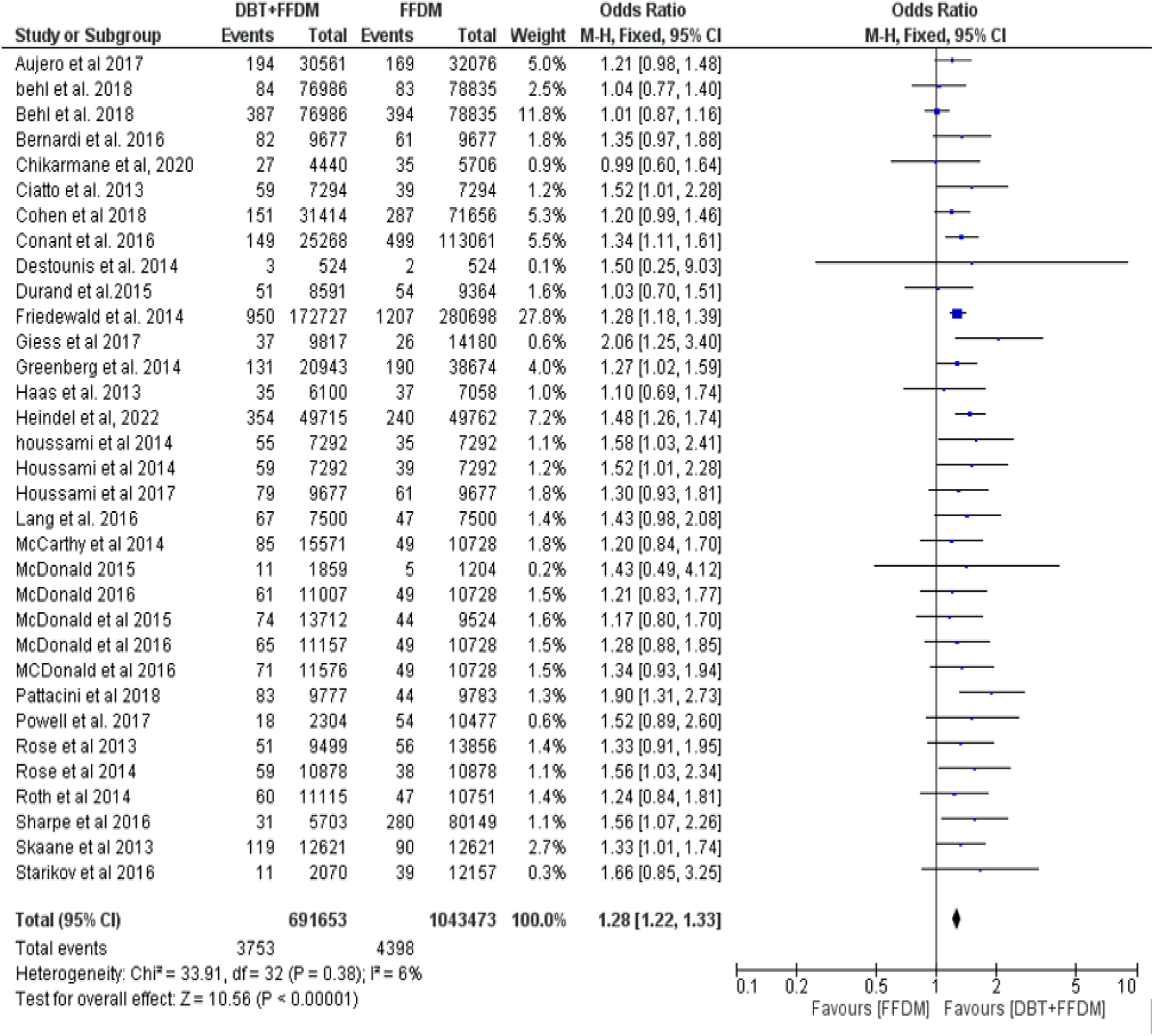
Forest chart of DBT+FFDM vs FFDM.

**Image 3:**
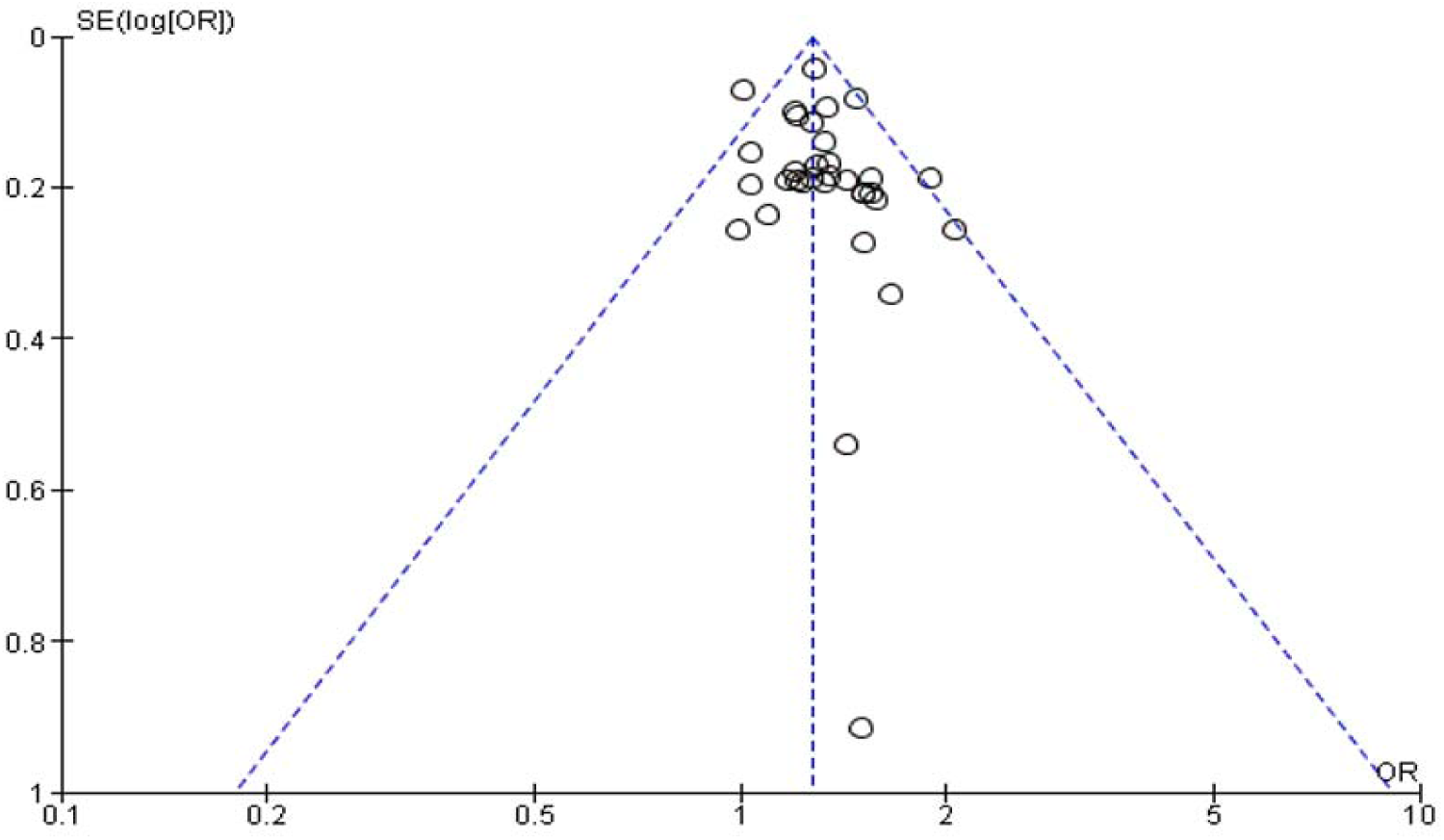
Funnel plot of DBT+FFDM vs FFDM.

### DBT vs FFDM

A total of 5 RCTS with 554,419 patients were selected for this study, Image 4 describes the forest charts and Image 5 describes the funnel plot. Here overall diagnostic odds ratio was 1.38 with a 95%CI [1.28 to 1.50]. here, it is also notable that an increase in sample size leads to an increase in the odds ratio for DBT. This can be seen in Alsheik et al 2019 and Winter et al 2020.

**Image 4:**
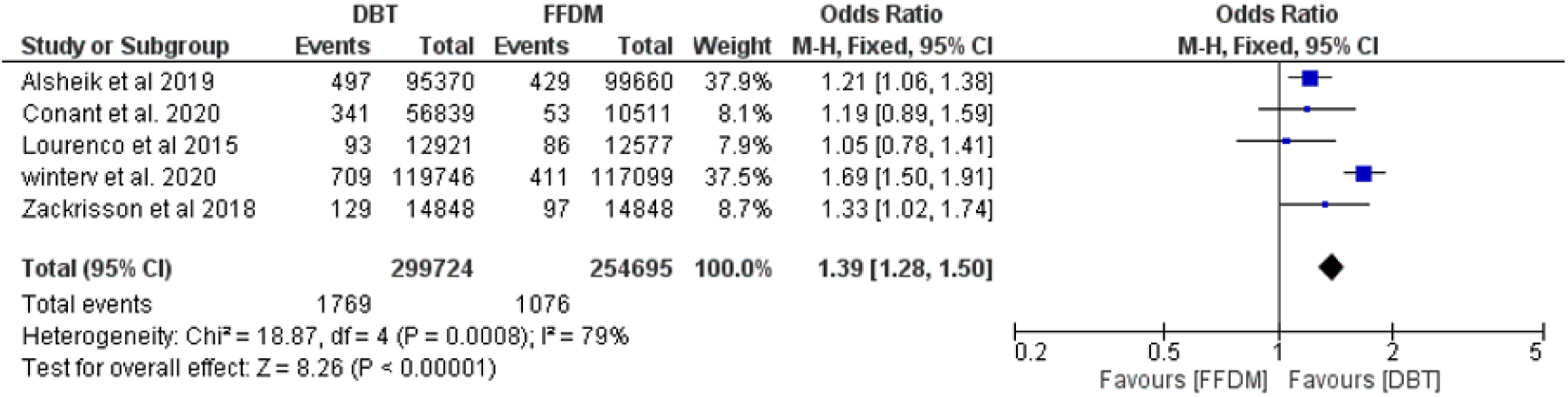
Forest chart of DBT vs FFDM.

**Image 5:**
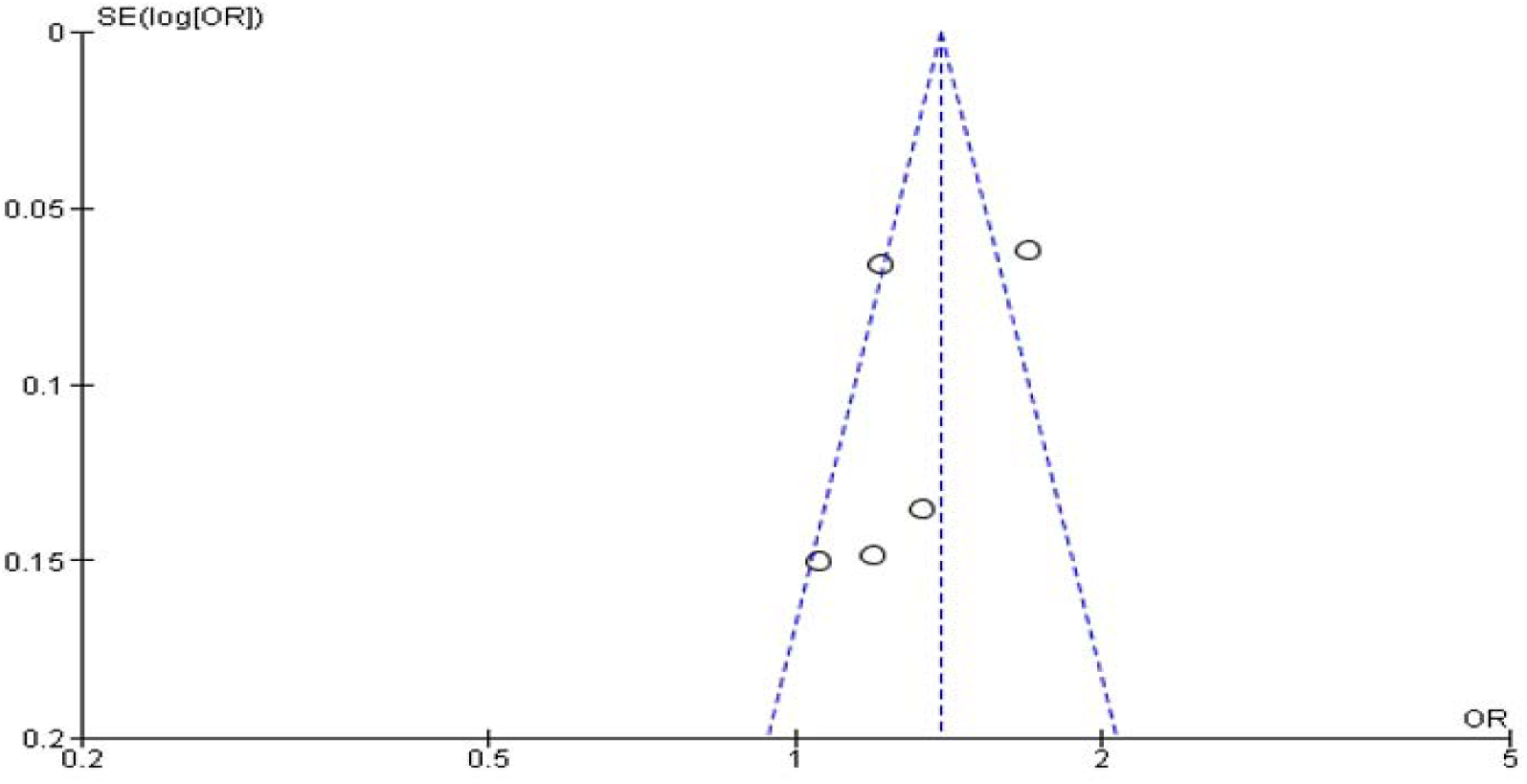
Funnel plot of DBT vs FFDM.

#### Decrease in the recall rate

Patients within the screening with BI-RADS 0, 3, 4, or 5 were recalled for further investigation and the recall rate was calculated.

Table 2 and Table 3 show a decrease in recall rate for DBT+FFDM and DBT respectively. For DBT+FFDM vs FFDM, the decrease in recall rate is 2.43% overall. For DBT vs FFDM, the average decrease in recall rate was 1.70%.

**Table 2:**
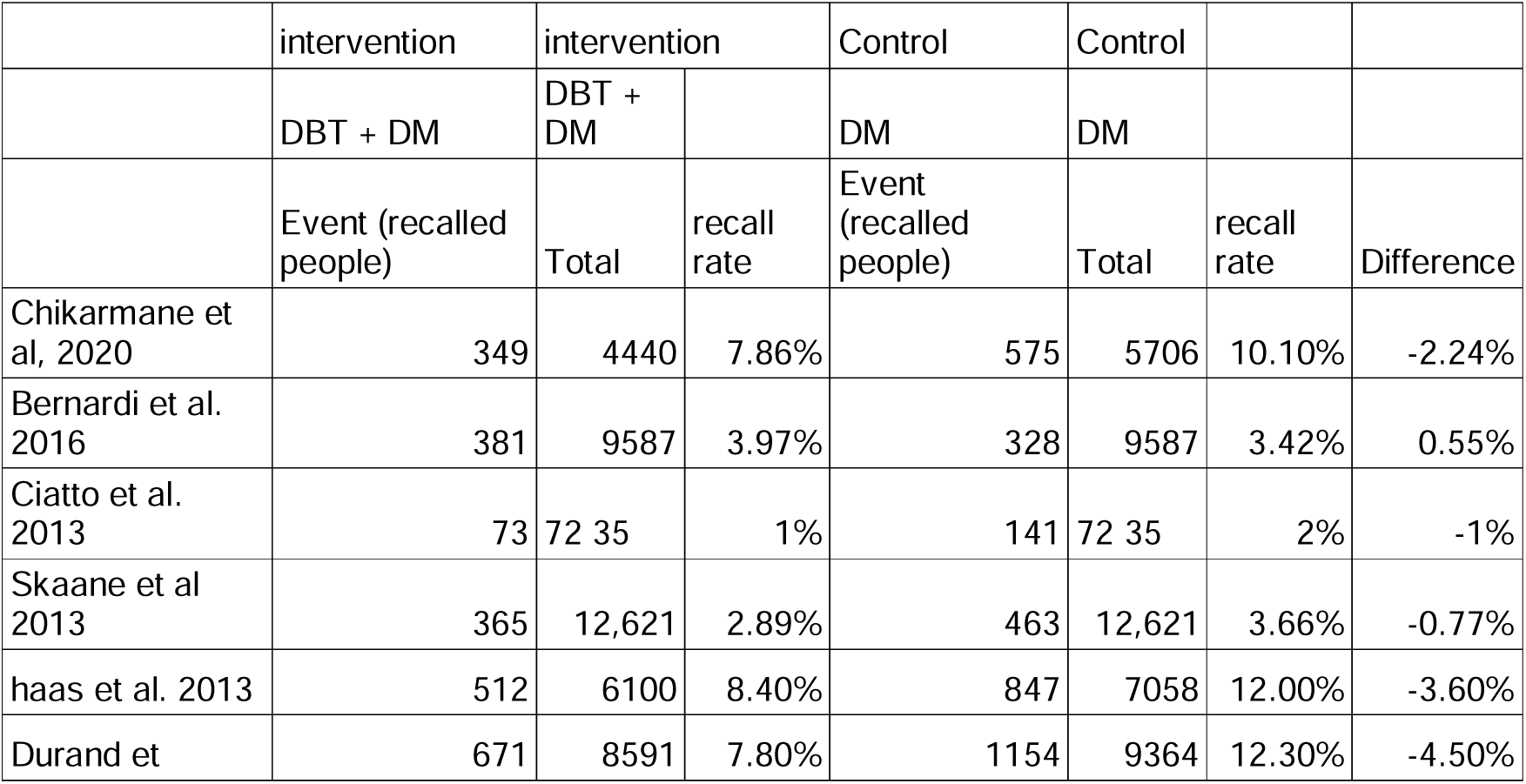
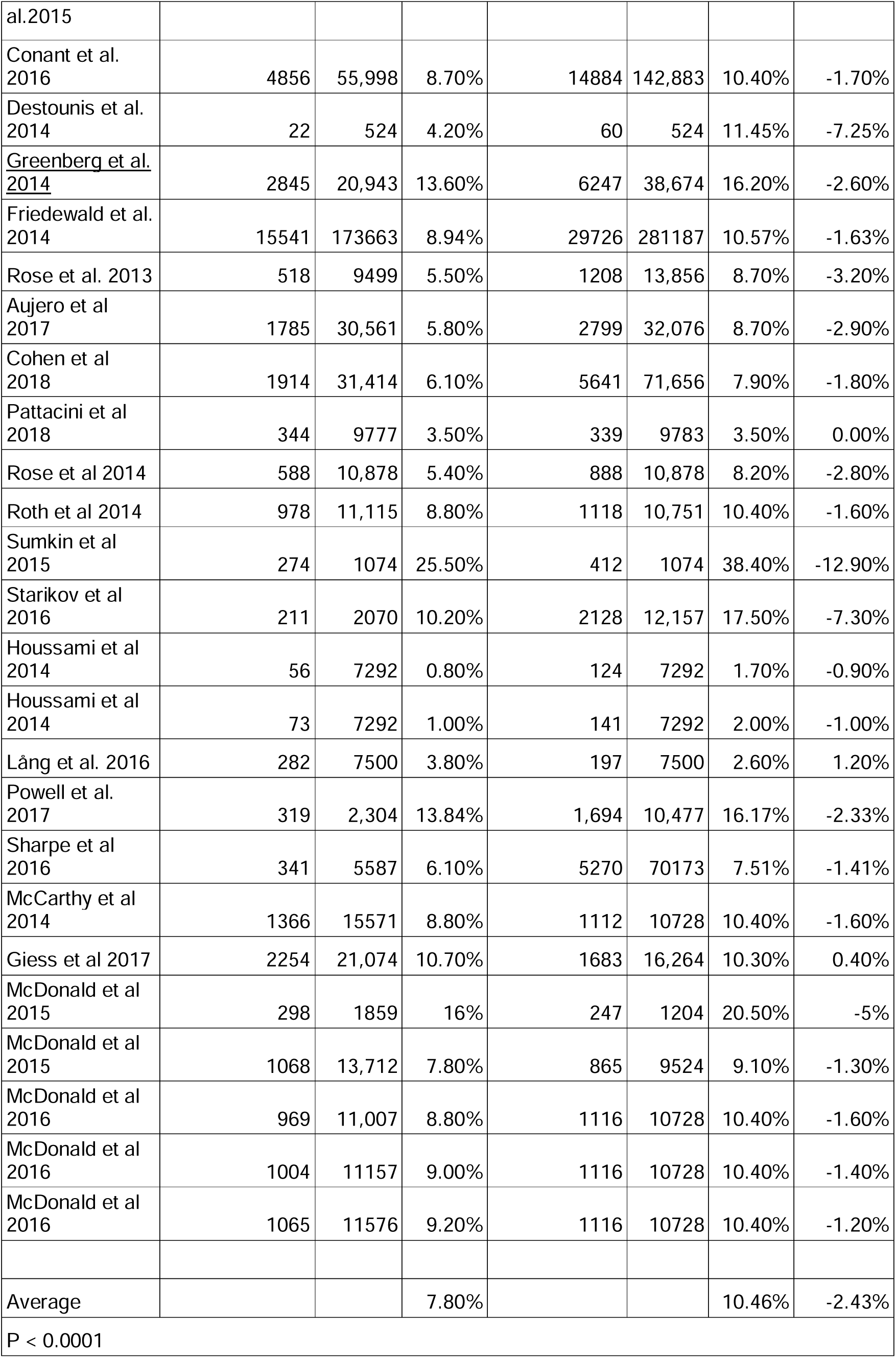
Decrease in recall for DBT+FFDM vs FFDM.

**Table 3:**
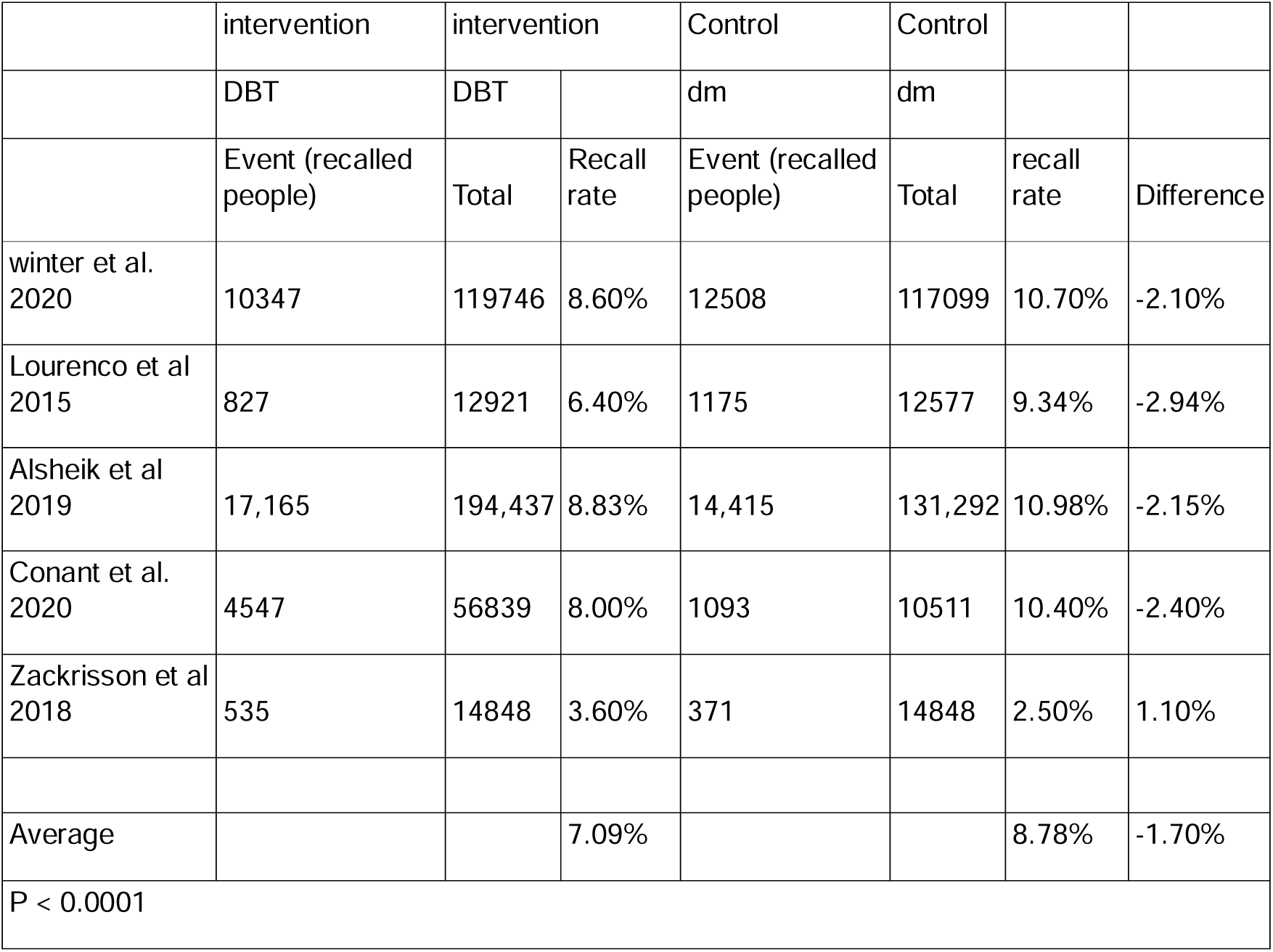
Decrease in Recall Rate for DBT vs FFDM.

#### Increase in Cancer detection rate

Table 4 and Table 5 show an increase in cancer detection rate for DBT+FFDM and DBT respectively. The average increase in cancer detection rate was 1.583 for DBT+FFDM and for DBT it was 1.4.

**Table 4:**
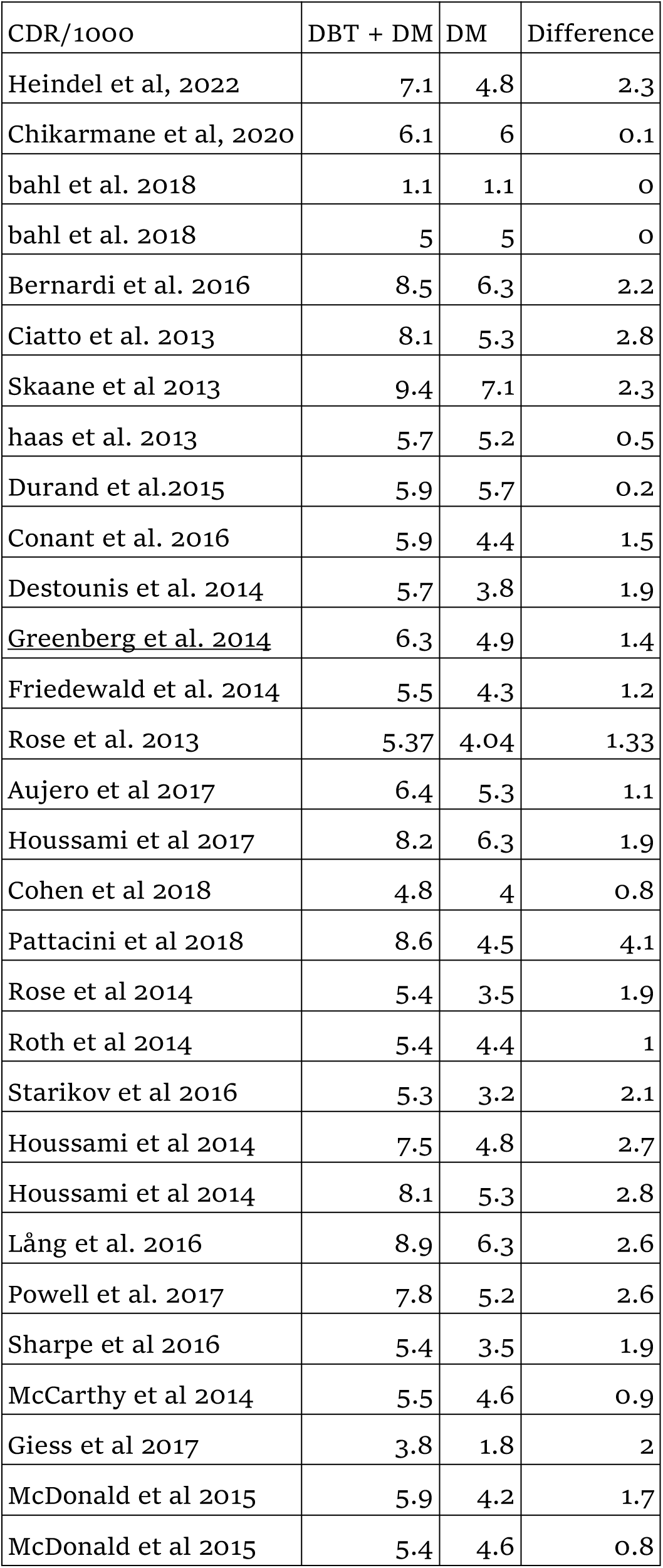
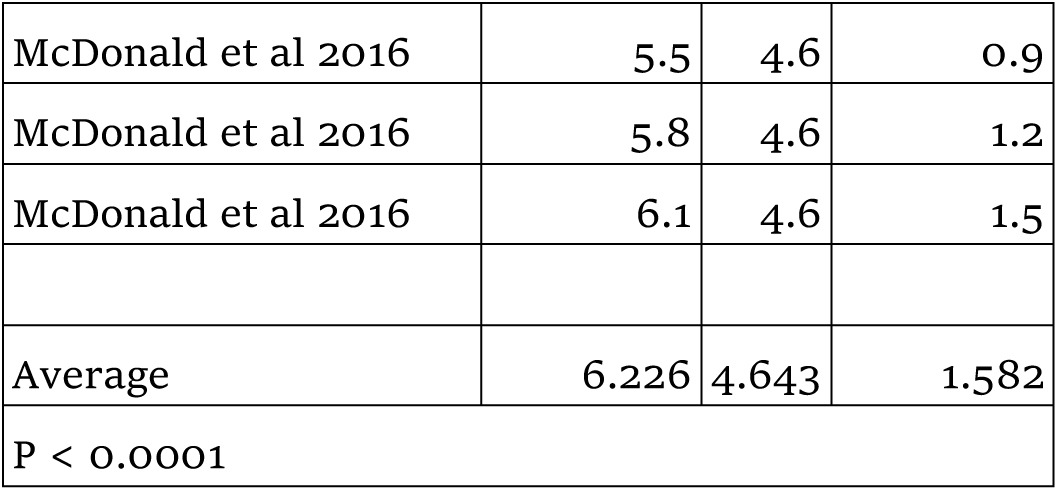
Cancer detection rate for DBT+FFDM vs FFDM.

**Table 5:**
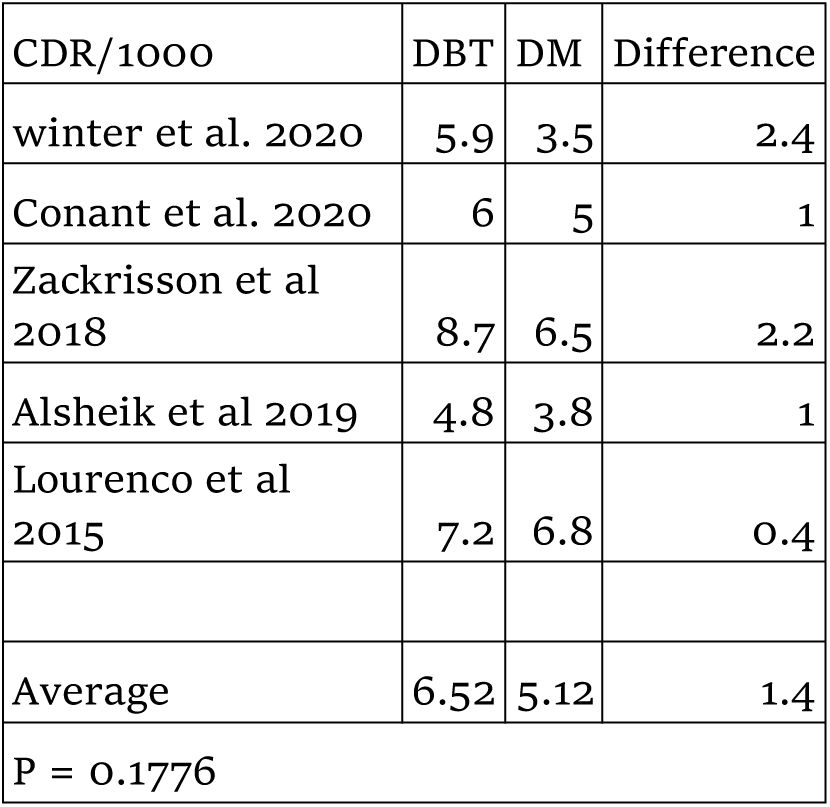
Cancer detection rate for DBT vs FFDM.

## Bias Study

**Table 6:** Risk of bias and applicability concern.

|  | Author<br>Name and<br>Year | Patient<br>Selection | Index test | Reference<br>standard | Flow and<br>timing |  | Patient<br>selection | Index test | Reference<br>standard |
| --- | --- | --- | --- | --- | --- | --- | --- | --- | --- |
| 1 | Alsheik<br>et al 2019 | low | low | High | high |  | high | low | low |
| 2 | Aujero<br>et al 2017 | Low | low | Low | low |  | Low | Low | low |
| 3 | Bahl<br>et al 2018 | low | low | low | high |  | low | low | low |
| 4 | Bernardi<br>et al 2016 | unclear | low | Low | high |  | low | low | low |
| 5 | Chikarman<br>e<br>et al 2020 | low | low | low | unclear |  | unclear | low | low |
| 6 | Ciatto<br>et al 2013 | Low | Unclear | Low | Low |  | low | low | Low |
| 7 | Cohen<br>et al 2018 | Low | Low | Unclear | Low |  | Low | Low | Low |
| 8 | Conant<br>et al 2016 | Low | Unclear | Low | Low |  | Low | unclear | Low |
| 9 | Conant<br>et al 2020 | Low | Low | Low | unclear |  | Unclear | Low | low |
| 10 | Destounis<br>et al 2014 | Low | High | unclear | Low |  | Low | Low | unclear |
| 11 | Durand<br>et al 2015 | unclear | Low | Low | unclear |  | Low | Low | low |
| 12 | Friedewald<br>et al 2014 | Low | Unclear | Low | Low |  | Low | unclear | low |
| 13 | Giess<br>et al 2017 | low | Low | High | high |  | high | low | low |
| 14 | Greenberg<br>et al 2014 | low | Low | unclear | unclear |  | low | low | low |
| 15 | Haas<br>et al 2013 | Low | Low | Low | Low |  | Unclear | Low | unclear |
| 16 | Heindel<br>et al 2022 | Low | Low | Low | low |  | Low | Unclear | low |
| 17 | Houssami<br>et al 2014 | Low | Low | Low | Low |  | Unclear | Low | unclear |
| 18 | Houssami<br>et al 2017 | unclear | Low | unclear | low |  | Low | Low | low |
| 19 | Lang<br>et al 2016 | Low | low | Low | unclear |  | Low | Low | unclear |
| 20 | Lourenco<br>et al 2015 | Low | Unclear | Low | Low |  | low | high | Low |
| 21 | McCarthy<br>et al 2014 | Low | unclear | high | Low |  | Low | Low | unclear |
| 22 | McDonald<br>et al 2015 | Low | High | Low | unclear |  | Low | Low | low |
| 23 | McDonald<br>et al 2016 | Low | Low | Low | Low |  | Unclear | Low | unclear |
| 24 | Pattacini<br>et al 2018 | unclear | low | Low | Low |  | Low | high | unclear |
| 25 | Powell<br>et al 2017 | low | Low | unclear | high |  | Low | Low | low |
| 26 | Rose<br>et al 2013 | Low | Unclear | Low | Low |  | low | low | Low |
| 27 | Rose<br>et al 2014 | Low | Unclear | Low | Low |  | low | low | Low |
| 28 | Roth<br>et al 2014 | unclear | low | Low | low |  | Low | Low | low |
| 29 | Sharpe<br>et al 2016 | Low | low | high | Low |  | Low | Low | low |
| 30 | Skaane<br>et al 2013 | Low | low | Low | low |  | Low | unclear | low |
| 31 | Starikov<br>et al 2016 | Low | low | unclear | low |  | Low | Low | low |
| 32 | Sumkin et al 2015 | low | low | high | low |  | high | low | low |
| 33 | Winter et al 2020 | low | low | low | high |  | low | unclear | low |
| 34 | Zackrisson et al 2018 | low | low | low | low |  | low | unclear | low |

### Publication Bias

The summary of publication bias for DBT+FFDM vs FFDM and DBT vs FFDM is shown in (Image 6). For the publication bias, In, patient selection, bias was low for 29 studies and unclear for 5. In the index test, bias was low in 25 studies, unclear in 7, and high in 2 studies. While for the reference standard, the bias was low in 23, unclear in 6, and high in 5 studies. The flow and timing bias was low in 22, unclear in 6, and high in 6. The applicability concerns bias in patient selection were low in 26, unclear in 5, and high in 3 studies. The index test bias was low in 26, unclear in 6, and high in 2. The reference standard bias was low in 27, and unclear in 7.

**Image 6:**
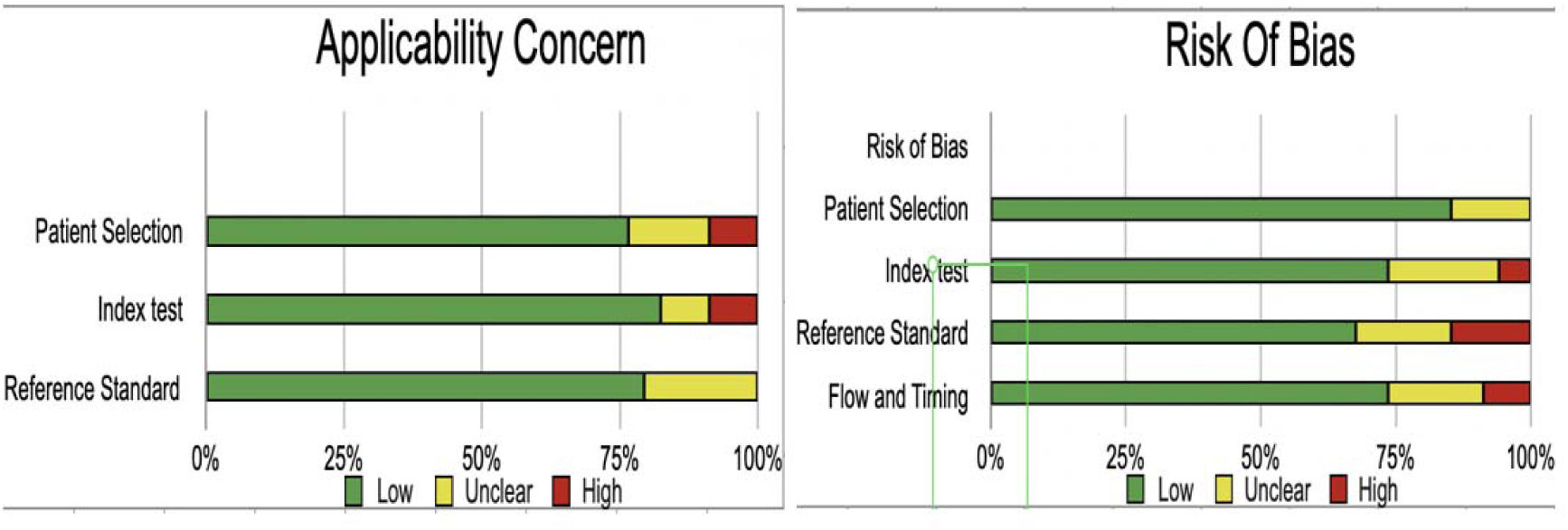
Risk of bias and Applicability Concern.

## Discussion

To present the best available evidence to help clinicians with decision-making, this study was conducted as a meta-analysis. The primary aim of this study was to evaluate the increase in cancer detection rate[CDR] and decrease in recall rate[RR] for 3D+2D vs 2D and 3D vs 2D mammography. The cancer detection rate was calculated per 1000 population screened, and every patient with BI-RADS category for highly suggestive cat 5 and cat 6. The results show, with a mild to moderate quality of evidence, that implementing DBT plus digital or synthetic mammography in population-based breast cancer screening increases overall breast cancer detection rates, and decreases recall rate.

For Lourenco et al 2015, the screening examinations performed during this study are taken as the total population and not the actual population as the data is provided in such a way in the paper.

Multiple studies have shown that DBT can substitute mammographic spot compression views for the evaluation of noncalcified lesions. The decrease in RR for asymmetries and focal asymmetries with DBT is related to improved characterization of overlapping breast tissue as a benign finding, which would reduce RR for summation of overlapping tissue. It may also be related to improved visualization of mass margins. According to Zuley et al 2013, By using tomosynthesis in the screening, radiologists were able to find fewer benign masses as BIRADS category 3, 4, or 5, without loss of much sensitivity, and malignant lesions were found as highly suggestive of malignancy (BI-RADS category 5), and with the help of tomosynthesis, radiologists were able to differentiate between malignant and benign lesions better. [47]–[50]

In this study, The CDR is identified with an average of 1.28 for 3D+2D vs 2D and 1.39 for 3D vs 2D study design. It has been seen in the result that the incorporation of DBT into conventional 2D mammography does increase the CDR. However, in comparison to radiation exposure, 3D+2D is almost double the exposure than 2D alone. This may theoretically limit the application of DBT with DM.[5]

As the usage of DBT in screening is limited, it may be very useful for the detection of benign Architectural distortions and Assessment of symptomatic women and work-up of screening-detected suspicious findings as lumps are identified by their shape, size margins, density, and content. The shape of a lesion is often better defined at DBT and DBT+FFDM than FFDM alone.[50], [51]

However, DBT-guided FNAC, DBT-guided vacuum-assisted breast biopsy, Contrast-media-enhanced DBT, and Fusion imaging like DBT with other 3D modalities such as 3D-automated breast ultrasound, radionuclide imaging, or even MRI is a promising research approach.[5], [52]

With some of these limitations of the radiation exposure from DBT, there is a new opportunity for using 2D synthesised-3D mammography, which may show less radiation exposure and almost the same or more CDR than DBT+FFDM.

## Limitations

This study has not divided the study population into sub-age groups and also some papers only involved BIRADS cat 0 in RR. For the diagnosis and incorporation into the routine screening, DBT requires more time and study and the equipment is costlier.

## Conclusion

As shown above, DBT+FFDM and DBT do outweigh FFDM alone and must be used with precaution and proper understanding of reading it as it can identify more cancers and can separate benign from malignancy better for BI-RADS 1,2 and highly suggestive of malignancy cat 5 and cat 6. In clinical practice, the use of tomosynthesis is likely to provide a lower recall rate, an increase in cancer detection rate, fewer screening examinations, fewer short-interval follow-up studies, and fewer biopsies for patients with benign lesions. Thus, relieving anxiety and resources of the patients and hospitals. Using this technology, patients can benefit more than its side effects. For hospitals where the patient load is very high and facilities that provide mass population screening and community diagnosis, these modalities can be implemented for the betterment of the population.

## Data Availability

All data produced in the present work are contained in the manuscript

